# Acute Cardiovascular and Electrocardiographic Effects of Nicotine Pouches: A study protocol for a Randomized, Double-Blind, Placebo-Controlled Crossover Trial (NICOTUNE STUDY)

**DOI:** 10.64898/2026.08.29.26361726

**Authors:** Elvin Khodi Babaroudi, Maria Hang Xuan Pham, Ingrid Try Lenz, Emilie Louisa Renner Melgaard, Jens Dahlgaard Hove, Johannes Grand, Ekim Seven

**Author notes:** **Corresponding author:** Dr. Elvin Khodi Babaroudi, MD. Department of Cardiology, Copenhagen University Hospital - Amager and Hvidovre Hospital. Kettegård Alle 30, 2650, Hvidovre, Denmark. **Email addresses for contributing authors:**.

## Abstract

**Introduction:** Nicotine Pouches are increasingly used as a smokeless alternative to cigarettes and other nicotine products, yet their acute cardiovascular effects remain poorly documented. While nicotine’s impact on heart rate and electrocardiogram (ECG) parameters is well-documented in smoking, no trials have evaluated these effects specifically for nicotine pouches.

**Methods:** This study is a single-center, double-blind, placebo-controlled, crossover trial which will include 20 healthy adult nicotine users. Participants will undergo three sessions, receiving either a placebo, 6 mg, or 14 mg nicotine pouch in random order. Heart rate obtained by an ECG and various other ECG parameters, vital signs, and subjective symptoms will be measured at baseline, and multiple time points over 30 minutes.

**Conclusions:** This study aims to determine whether nicotine pouches cause acute changes in heart rate, ECG parameters, vital signs, and self-reported symptoms. We hypothesize that higher nicotine pouch does will lead to measurable increases in heart rate and other autonomic effects compared to placebo.

**Trial registration:** The study has been approved by The Committees on Health Research Ethics in the Capital Region of Denmark (H-25018523). The study was registered with Clinicaltrial.org on 2025.07.09 (Identifier: NCT07057440).

## Introduction and Background

Nicotine pouches are smokeless, tobacco-free products designed to deliver nicotine through oral transmucosal absorption. They have gained popularity in recent years as an alternative to traditional smoking, vaping, and other nicotine replacement therapies like gums and patches, especially among teenagers and young adults.^1^ From 2016 to 2020, sales in the United States of America increased by 305%, and the market is expected to approach a value of nearly $33 billion by 2026. ^2,3^ Nicotine pouches are discreet, portable, and available in a range of nicotine concentrations, making them particularly appealing to individuals seeking to reduce smoking or avoid the stigma associated with other forms of nicotine use.^1^

Nicotine is a known stimulant of the autonomic nervous system, with well-documented acute effects on the cardiovascular system. These include increased heart rate, elevated blood pressure, and changes in electrocardiographic (ECG) parameters, such as QT interval prolongation and potential arrhythmogenic effects.^4–9^ However, these effects have predominantly been studied in relation to cigarette smoking and other nicotine-delivery systems. Nicotine pouches represent a distinct mode of nicotine administration through the oral mucosa, and previous studies have demonstrated variation in nicotine delivery and pharmacokinetic profiles across pouch products and nicotine strengths.^10–16^

Although several clinical studies have characterized the pharmacokinetics and subjective effects of nicotine pouches ^10–16^ only a limited number have assessed acute cardiovascular responses ^10,11,14^ and systematic evaluation of electrocardiographic changes has been limited. Thus, while the chronotropic effects of nicotine itself are well established, the magnitude and pattern of acute cardiovascular and electrocardiographic changes following nicotine pouch use remain incompletely characterized.

Accordingly, this study is designed to investigate the acute effects of nicotine pouches on heart rate and other ECG parameters in adults without known cardiovascular disease. By comparing 6 mg and 14 mg nicotine pouches with placebo under standardized crossover conditions, the study aims to characterize the cardiovascular response associated with nicotine pouch use. We hypothesize that nicotine pouch use will increase resting heart rate compared with placebo, with a greater increase following the 14 mg pouch. Secondary hypotheses include changes in ECG parameters, vital signs, and subjective symptoms.

## Methods

### Study Design and Setting

This study is a single-center, double-blind, placebo-controlled crossover trial designed to assess the acute effects of nicotine pouches on cardiac electrophysiology in healthy adults. The trial will be conducted at the Department of Cardiology, Copenhagen University Hospital, Amager-Hvidovre, located in the Capital Region of Denmark. The study will take place in a controlled clinical setting during the summer and autumn of 2025 and is registered with ClinicalTrials.gov (Identifier: NCT07057440).

The study is an investigator-initiated trial initiated by Doctor Ekim Seven and Doctor Elvin Khodi Babaroudi. The remaining individuals of the steering group has provided valuable feedback regarding the design of the study protocol or helped with the execution of the study.

### Outcomes (Endpoints)

The outcomes of the study are listed in table 2. The primary outcome is the change in heart rate, measured in beats per minute from the 12-lead ECG, at 20 minutes after nicotine pouch administration compared with the session-specific baseline. The 20-minute time point was prespecified because it coincided with the end of the standardized pouch exposure period, which followed the manufacturer’s recommended duration of use. This time point was therefore selected to capture the cardiovascular response at the end of active pouch exposure. Pharmacokinetic studies of nicotine pouches have also reported peak plasma nicotine concentrations within approximately 20–30 minutes for some formulations. ^10,14^

**Table 1:**
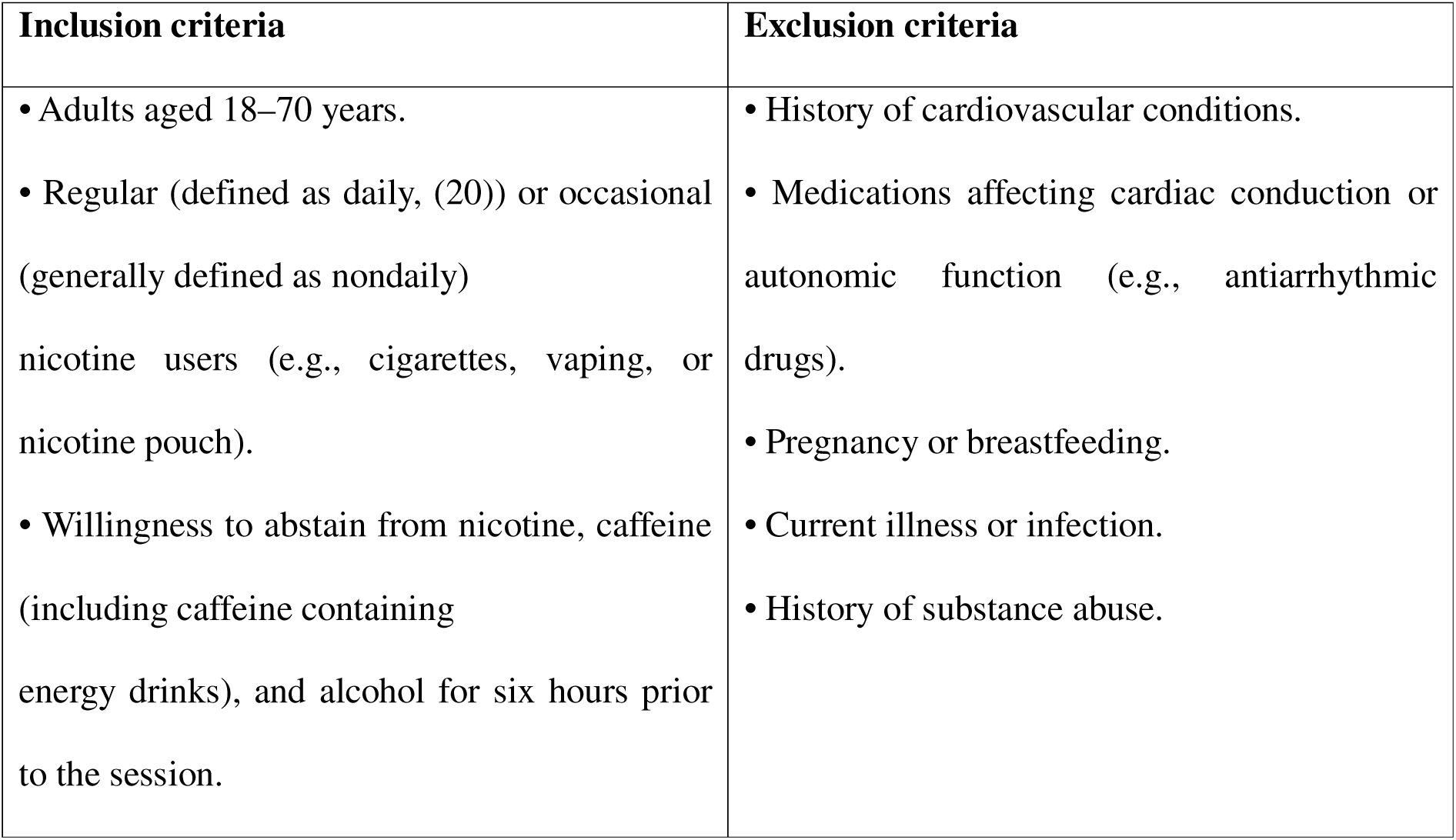
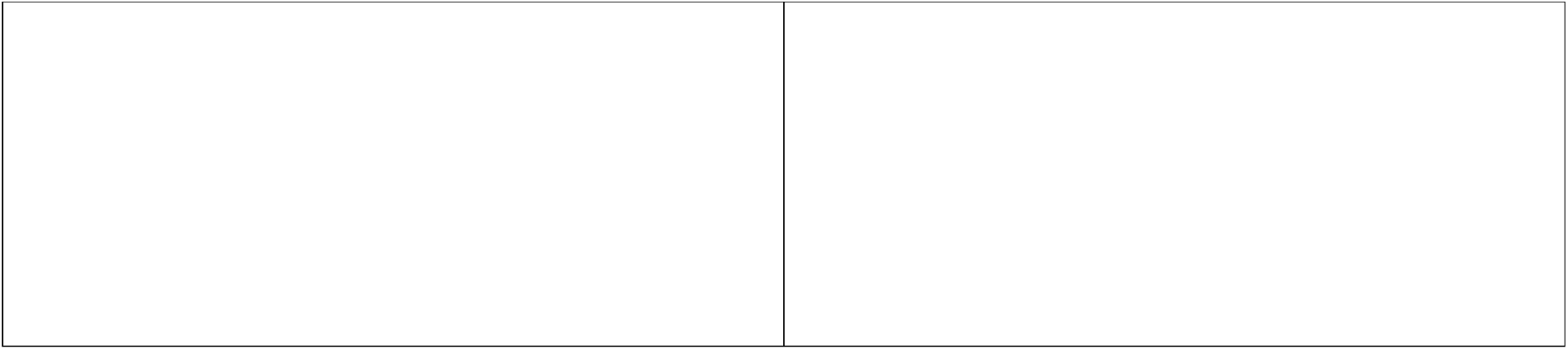
Eligibility criteria.

| Inclusion criteria | Exclusion criteria |
| --- | --- |
| • Adults aged 18–70 years.<br>• Regular (defined as daily, (20)) or occasional (generally defined as nondaily) nicotine users (e.g., cigarettes, vaping, or nicotine pouch).<br>• Willingness to abstain from nicotine, caffeine (including caffeine containing energy drinks), and alcohol for six hours prior to the session. | • History of cardiovascular conditions.<br>• Medications affecting cardiac conduction or autonomic function (e.g., antiarrhythmic drugs).<br>• Pregnancy or breastfeeding.<br>• Current illness or infection.<br>• History of substance abuse. |

**Table 2:**
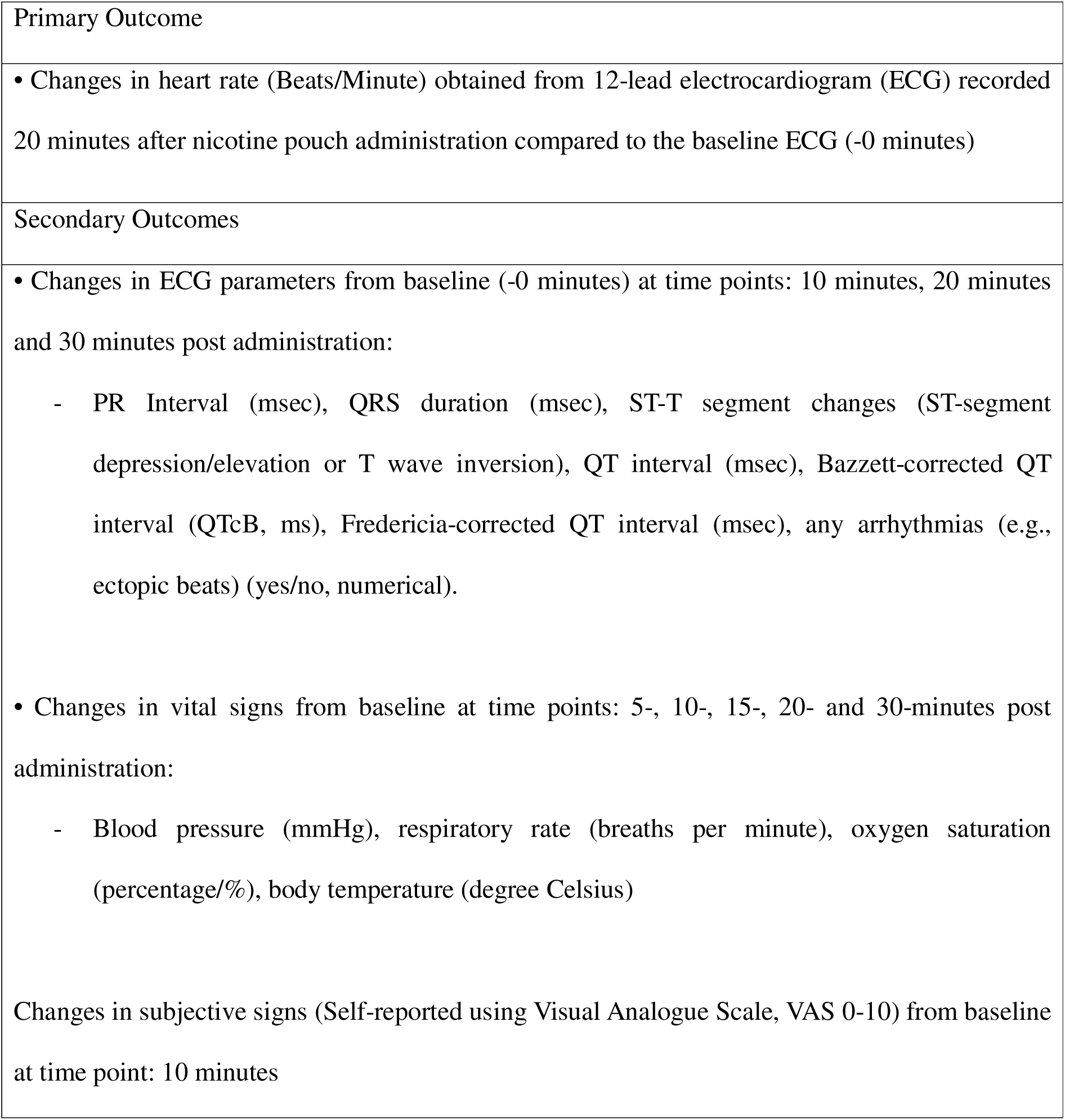
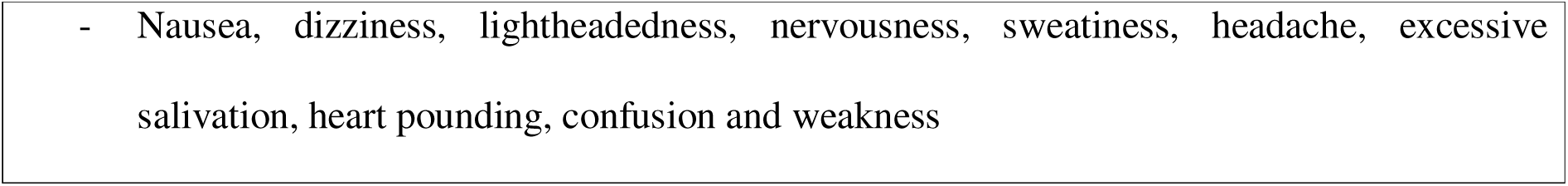
Primary and secondary outcomes.

Secondary outcomes include changes from baseline in prespecified ECG parameters at 10, 20, and 30 minutes after pouch administration, including PR interval, QRS duration, QT interval, QTcB, and QTcF. ST-segment and T wave abnormalities and ectopic beats will also be recorded.

Exploratory outcomes include changes in vital signs, including blood pressure, respiratory rate, oxygen saturation, and body temperature, as well as changes in self-reported nicotine-related symptoms assessed using a visual analogue scale (VAS). Ten predefined symptoms will be evaluated: nausea, dizziness, lightheadedness, nervousness, sweating, headache, excessive salivation, heart pounding, confusion, and weakness.

The sample size calculation is based exclusively on the primary heart rate outcome. Secondary and exploratory outcomes are included to characterize the broader acute cardiovascular response to nicotine pouch exposure and are not individually powered for confirmatory hypothesis testing.

### Participants and Eligibility Criteria

Participants will be considered eligible if they meet the following criteria.

1. Adults aged 18-70 years,
2. regular (defined as daily or occasional)^17^ nicotine users (e.g., cigarettes, vaping, or nicotine pouch),
3. Willingness to abstain from nicotine-containing products, caffeine (including caffeinated energy drinks), and alcohol for at least six hours before each study session (Table 1).^18^

Participants will be excluded if they meet any of the following criteria.

1. History of cardiovascular conditions,
2. medications affecting cardiac conduction or autonomic function (e.g., antiarrhythmic drugs),
3. pregnancy or breastfeeding,
4. current illness or infection,
5. history of substance abuse (Table 1).

### Study interventions

#### Interventions

Each participant will complete three study sessions on separate days, with a washout period of at least 24 hours between consecutive sessions. All study sessions will be conducted during the morning hours between 08:00 and 12:00 to standardize the timing of assessments and limit potential variation related to time of day. At each session, participants will receive one of three interventions in randomized order: a placebo pouch containing no nicotine, a 6 mg nicotine pouch, or a 14 mg nicotine pouch. Each nicotine pouch will be of the same taste, peppermint. The nicotine conditions refer to the nominal nicotine content of the pouches. Plasma nicotine concentrations were not measured as part of the study protocol.

#### Screening and Baseline Assessment

Prior to the intervention, all participants will undergo a screening and baseline assessment to ensure eligibility and establish reference values. This includes the collection of detailed medical history and demographic information, including age and sex, as well as a detailed nicotine use history, including nicotine product type and frequency of use. Participants will also be asked to rate the ten most common side effects associated with nicotine products using a VAS ranging from 0-10.^19^ Anthropometric measurements such as weight, height, and body mass index (BMI) will be recorded. Additionally, resting vital signs including heart rate, (BP, respiratory rate, SAT, and body temperature will be measured under standardized conditions. Finally, a baseline 12-lead ECG will be performed to assess cardiac electrical activity prior to intervention (Table 3 and Figure 1).

**Figure 1:**
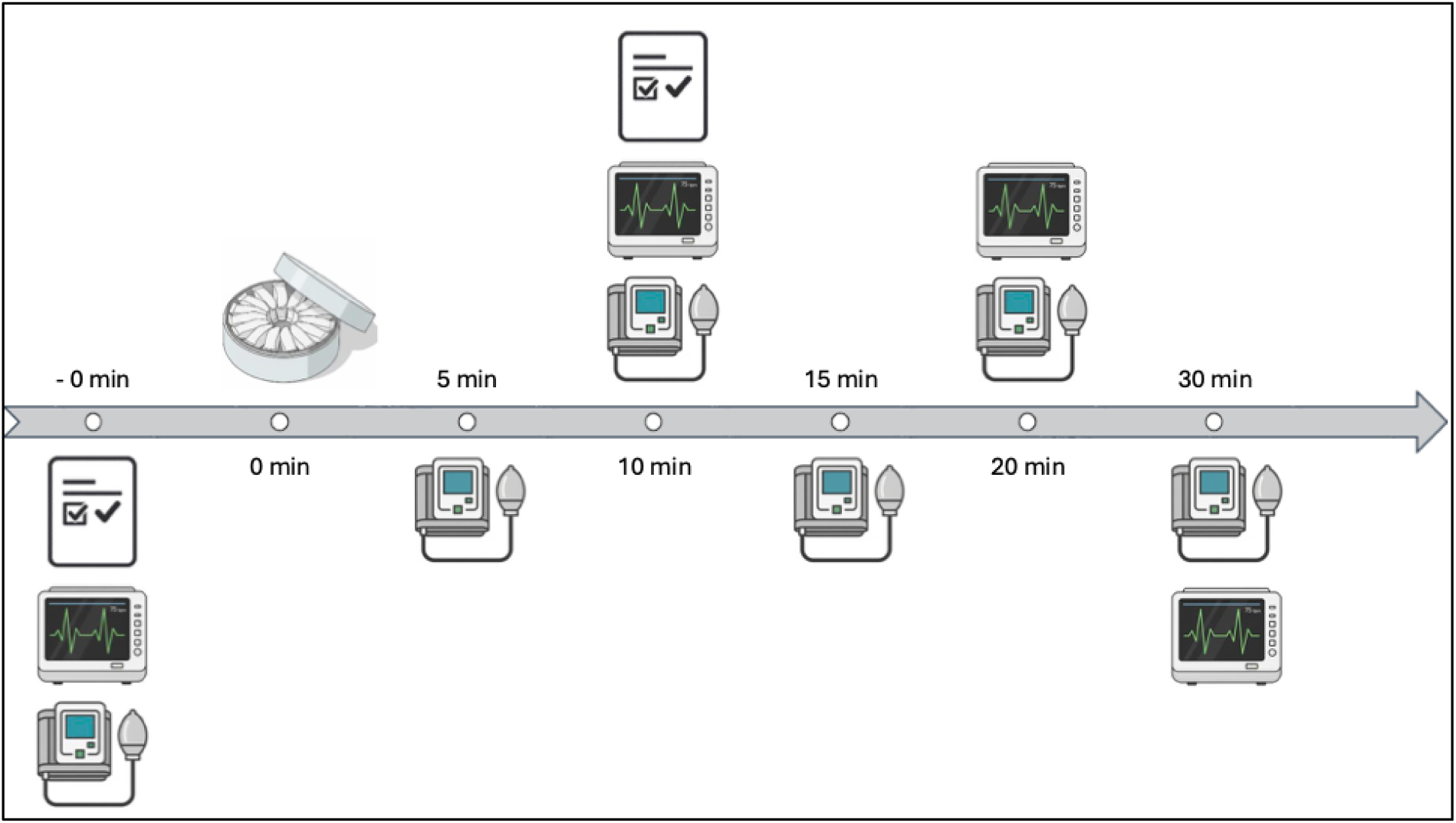
Intervention monitoring figure. -0 min: Participants will go through the resting phase with screening and baseline assessments, anthropometric examination, VAS screening of symptoms and an ECG. 0 min: The nicotine pouch will be given. 5 min: Anthropometric examination. 10 min: Anthropometric examination, VAS screening of symptoms and an ECG will be performed. 15 min: Anthropometric examination. 20 min: nicotine pouch will be taken out, anthropometric examination and an ECG will be performed. 30 min: Anthropometric examination and an ECG will be performed.

**Table 3:**
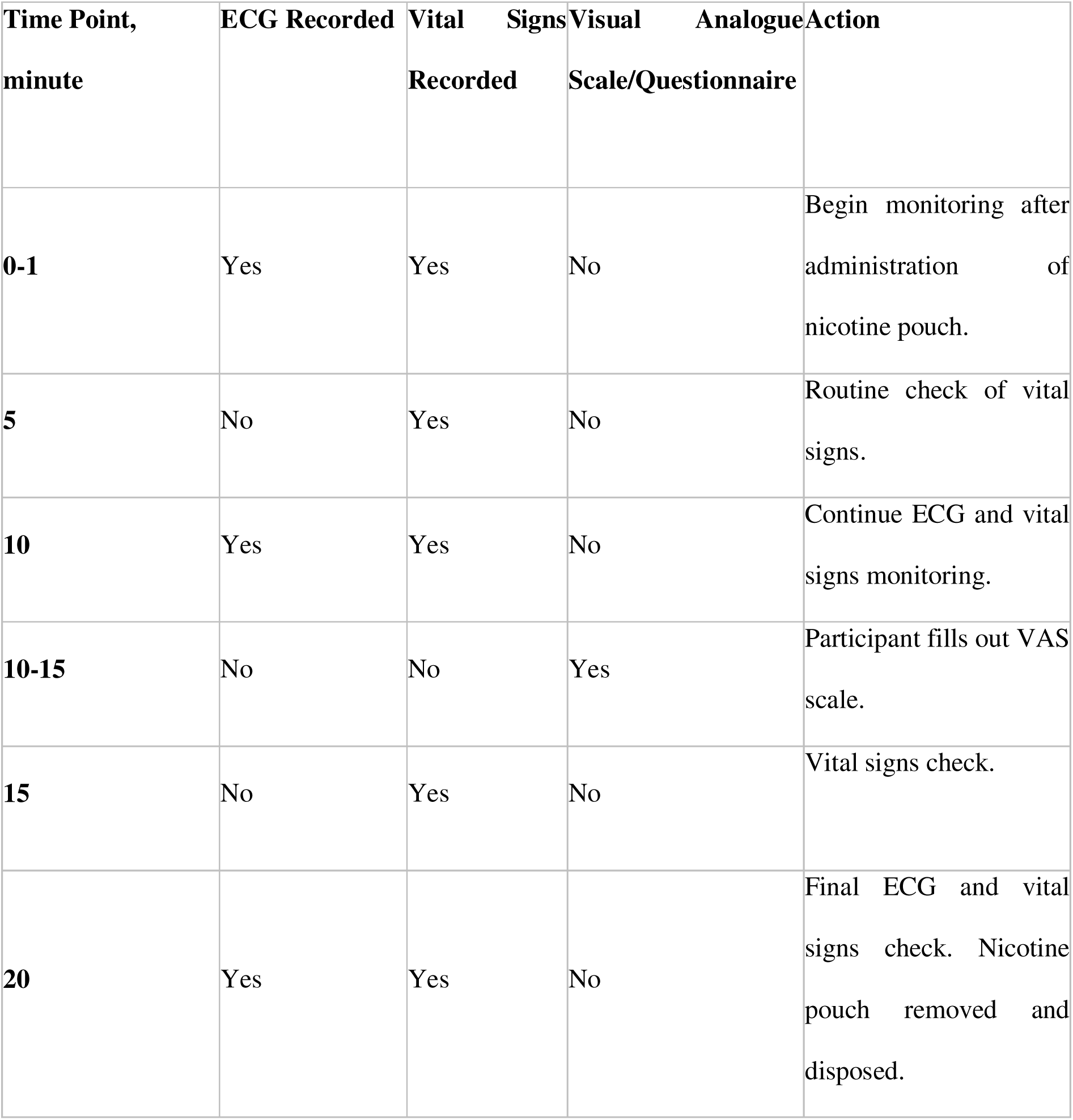
Intervention monitoring table.

#### Randomization and Blinding

Each type of nicotine pouch will be placed in a transparent tray without any labelling that could reveal the brand, manufacturer, or nicotine conditions of the product. To facilitate blinding while allowing the internal categorization, each tray will be marked on the top with a colored label, indicating the assigned nicotine condition category. Randomization and labelling will be handled by designated study coordinators who will not be involved in data collection, analysis, or outcome assessment, thereby maintaining double-blind conditions throughout the trial. For each of the three experimental sessions, one of the three pouches will be randomly assigned in a within-subject crossover manner.

#### Intervention Protocol

Each study session will be divided into three distinct phases: A resting phase, an exposure phase, and a post-exposure monitoring phase (Table 3 and Figure 1).

Upon arrival at the study site, participants will enter the first phase, a 20-minute resting period during which they will remain in a quiet, seated or supine position to establish baseline physiological conditions. During this time, screening and baseline assessments will be conducted, including medical history review, nicotine use documentation, anthropometric measurements, resting vital signs, and a baseline 12-lead ECG. In the end the administration of the assigned nicotine pouch will take place. Participants will receive a nicotine pouch from a pre-labelled tray; the tray label code and the participant’s ID number will be recorded to ensure traceability.

The exposure phase starts the moment the participants place the nicotine pouch between the upper lip and gum and held in the oral cavity for 20 minutes, following the standard usage instructions by the manufacturer. This phase consists of systematic collection of ECG data and vital signs. ECGs will be recorded at 10 and 20 minutes into the intervention. Vital signs (BP, heart rate, respiratory rate, SAT temperature) will be measured at 4 time points: After 5, 10, 15 and 20 minutes into the intervention. Participants will also report a follow-up of subjective symptoms using VAS after the midpoint ECG, approximately 10-15 minutes after administration of the nicotine pouch. At each prespecified time point, a standard 10-second 12-lead ECG will be recorded digitally with the participant in a resting supine position. Heart rate, RR interval, PR interval, QRS duration, and QT interval will be obtained from the automated ECG measurements. Heart rate-corrected QT intervals will be calculated using both Bazett’s formula (QTcB) and Fridericia’s formula (QTcF). Because nicotine exposure is expected to induce changes in heart rate, QTcF will be considered the more reliable measure for interpretation of ventricular repolarization, while QTcB will also be reported because of its widespread clinical use.^20^

All ECGs will be reviewed by two experienced investigators blinded to treatment allocation. ST-segment and T wave abnormalities and ectopic beats will be assessed manually, and discrepancies in ECG interpretation will be resolved by adjudication by a board-certified cardiologist.

The final phase, post-exposure phase, begins immediately after the 20-minute exposure period and continue for an additional 10 minutes following the removal of the nicotine pouch. During this phase, the participant remains in a supine and resting position. At the end of the phase a final 12-lead ECG will be recorded, and vital signs will be measured once more to complete the monitoring process.

#### Risk and Adverse Events

The intervention in this trial is anticipated to pose minimal risk to participants, as it involves the use of commercially available, approved, and widely used nicotine pouches. The expected side effects are limited to well-documented, acute effects of nicotine, including dizziness, headache, nausea, palpitations, sweating, increased salivation and local oral irritation in relation to the nicotine pouches.^21^

The participants of this study are either current or occasional nicotine users and are therefore presumed to have developed a certain degree of tolerance, the likelihood and severity of adverse effect are expected to be low. Additionally, risks associated with study monitoring procedures, such as ECG and BP measurements, are minimal with only mild or temporary discomfort.

To further reduce risk and ensure participant safety, the following precautionary measures will be implemented: Only healthy adults without known cardiovascular disease will be enrolled, inclusion is limited to nicotine users to avoid exposing nicotine naïve individuals and to mitigate the risk of initiating dependence, and lastly participants will be closely monitored throughout the study, and qualified medical personnel will be present to respond to any adverse events immediately.

### Sample size

The sample size calculation was based on the primary heart rate outcome and a paired within-participant comparison between an active nicotine condition and placebo at 20 minutes. The calculation assumed a mean paired difference of 3 beats per minute and a standard deviation of the paired differences of 3 beats per minute. With a two-sided significance level of 1% and 90% power, 19 participants were required. ^21^

The assumed standard deviation refers to the variability of the within-participant paired differences rather than to the variability of repeated heart rate measurements within an individual. The calculation was not intended to power an overall three-condition treatment effect, a formal acute response trend, or the secondary and exploratory outcomes. To allow for potential withdrawal or incomplete measurements, the study aims to include 20 participants.

### Data Collection and Management

The trial database/eCRF will be constructed using REDCap software hosted by the Capital Region of Denmark. All study data will be processed and stored confidentially in accordance with the Danish Data Protection Act and the General Data Protection Regulation. Access to study data will be restricted to authorized study personnel. The study will also be registered in the Capital Region of Denmark’s internal research directory.

### Statistical analysis

All analyses will be performed using R. Categorical variables will be summarized as counts and percentages, and continuous variables as mean with standard deviation or median with interquartile range, as appropriate. The primary endpoint will be analysed using a linear mixed-effects model appropriate for the crossover design. Treatment condition will be included as a fixed effect, with placebo as the reference condition, and participant will be included as a random intercept to account for the within-participant correlation across study sessions. Study period will be included as a fixed effect to account for potential period-related variation. Continuous secondary ECG and physiological outcomes will be analysed using analogous mixed-effects models where appropriate. Rare or categorical ECG findings, including ectopic beats and ST-T abnormalities, will primarily be summarized descriptively because the study is not powered for uncommon events. All statistical tests will be two-sided. Secondary and exploratory analyses will be interpreted cautiously given the number of comparisons performed.

## Discussion

This study aims to investigate whether nicotine pouches induce acute changes in ECG parameters in healthy adult volunteers. In addition, our study will assess acute effects on vital signs and self-reported symptoms associated with nicotine use. Although the autonomic effects of nicotine on heart rate and ECG parameters are well-established in the context of smoking, the cardiovascular impact of nicotine pouches remains largely unstudied. To our knowledge, this will be the first randomized, placebo-controlled trial to systematically evaluate acute ECG changes following nicotine pouch exposure.

Few published clinical trials have researched the pharmacokinetics of nicotine pouches.^10–16,22^ Only three studies investigated cardiovascular effect or side effects of nicotine pouch use.^10,11,14^ Chapman et al. found no changes in heart rate or BP after a 10 mg nicotine pouch, while Lunell et al. reported that a 10.5 bpm increase after 60 minutes with a 6 mg nicotine pouch. In contrast, Mallock et al. reported that 30 mg nicotine pouches increased heart rate by 25 bpm shortly after use, whereas 20 mg and 6 mg nicotine pouches had milder effects. However, none of the studies performed systematic ECG analysis of their participants.

A key strength of the study is its double-blind, placebo-controlled, crossover design, which minimizes bias and allows each participant to serve as their own control. This design increases internal validity and statistical power, even with a relatively small sample size. Furthermore, the study’s use of objective, quantitative endpoints, including standard 12-lead ECG recordings, vital signs, and VAS scores, ensures a rigorous and reproducible methodology. The trial setting with a controlled clinical environment also allows for consistent monitoring and adherence to protocol.

This study presents few limitations that should be acknowledged. First, the study sample is limited to healthy adults aged 18-70 years old with no known cardiovascular disease. While this increases participant safety, it limits generalizability to populations with pre-existing cardiac conditions, who may respond differently to acute nicotine pouches. Although the study is powered to detect changes in heart rate, it may not be sufficiently powered to evaluate rarer but clinically important arrhythmic events. Furthermore, the use of intermittent 10-second ECG recordings rather than continuous electrocardiographic monitoring limits the detection of transient arrhythmias or other ECG abnormalities occurring between the prespecified recording time points. Sex-specific differences in cardiovascular responses to nicotine cannot be excluded. The study was not designed or powered to assess treatment-by-sex interactions, and any potential sex-related differences should therefore be investigated in larger studies. Participants were required to abstain from nicotine-containing products for at least six hours before each study session. Given the pharmacokinetics of nicotine and the heterogeneity in habitual nicotine use, residual nicotine exposure cannot be completely excluded, and differences in nicotine tolerance may have influenced the magnitude of the acute cardiovascular response.^18^ The use of session-specific baseline measurements and the within-participant crossover design reduces, but does not eliminate, this potential source of variability. Plasma nicotine concentrations were not measured. Consequently, nicotine pouch condition was used as a proxy for nicotine exposure, and interindividual differences in nicotine extraction and systemic absorption could not be quantified. The study therefore cannot assess concentration-response relationships or directly relate the observed cardiovascular and electrocardiographic changes to measured systemic nicotine exposure. The monitoring period was limited to 30 minutes after pouch placement and was therefore designed to characterize the early cardiovascular response rather than the complete pharmacodynamic time course. Although some nicotine pouch formulations reach peak plasma nicotine concentrations within approximately 20-30 minutes, later peaks have also been reported and appear to depend, among other factors, on product characteristics and duration of pouch use.^10,14,15^ Consequently, cardiovascular effects occurring beyond the 30-minute observation period may not have been captured. The study population was heterogeneous with respect to habitual nicotine exposure, including users of different nicotine products and participants with varying frequencies of nicotine use. Although nicotine use history was recorded, validated measures of nicotine dependence and withdrawal were not included in the study protocol.

Differences in habitual exposure, nicotine tolerance, and withdrawal status may therefore have contributed to interindividual variability in cardiovascular and subjective responses. Pre-session abstinence from nicotine was not biochemically verified, and no expired-air carbon monoxide or plasma nicotine measurements were obtained. Consequently, complete adherence to the six-hour abstinence requirement cannot be objectively confirmed, and residual nicotine exposure at the start of a study session cannot be excluded. Formal assessment of blinding success was not included in the study protocol. Because nicotine may produce perceptible physiological or subjective effects, particularly at higher pouch strengths, functional unblinding of participants cannot be excluded.

If the trial confirms an acute alteration on ECG parameters, symptoms and basic vital signs these findings may have several implications for public health and consumer behavior, clinical practice and future research. From a public health perspective, it will clarify whether nicotine pouches exert significant acute cardiovascular and ECG effects, information that can guide risk communication and sager consumer choices, especially for individuals with heart conditions. In clinical practice, the study may inform future nicotine replacement therapy strategies by characterizing dose-response relationships. From a regulatory standpoint, findings could influence guidelines on nicotine content, labelling, and product warnings. Finally, in the scientific context, the results will contribute to the field of cardiovascular research and serve as a comparative reference point for other nicotine delivery systems.

## Conclusion

This trial aims to investigate the acute cardiovascular effects of nicotine pouches in healthy adults. By systematically comparing low, high, and placebo nicotine pouches, this study will provide the first controlled data on ECG changes, vital signs, and subjective side effect associated with nicotine pouch use. The findings are expected to contribute significantly to the scientific understanding of nicotine pouch safety, inform clinical practice, and guide future a regulatory framework.

## Data Availability

The datasets generated during the study will not be publicly available because of applicable data protection requirements but may be made available from the corresponding author upon reasonable request and subject to appropriate ethical and legal approvals.

## Declarations

### Ethics approval and consent to participate

This study will be conducted in accordance with the ethical principles outlined in the Declaration of Helsinki, ensuring respect for autonomy, safety and data integrity. The study has been approved by The Committees on Health Research Ethics in the Capital Region of Denmark (H-25018523). Written informed consent will be obtained from all participants prior to enrolment. Participants will receive both oral and written information about the study and will be informed of their right to withdraw at any time without consequences.

### Consent for publication

Not applicable. No individual person-identifiable data will be included in any publication arising from this study.

### Authors’ contributions

EK performed the primary study coordination, participant recruitment, coordination and scheduling of the study visits and data collection. EK will later lead the statisticial analyses and the drafting of the main manuscript. MHXP, ITL and ELRM contributed to participant recruitment and data acquisition. JDH and JG contributed to the study design, methodological development, and critical revision of the manuscript for important intellectual content. ES was the principal investigator of the study and, together with EK, played a central role in the development of the study protocol. ES and EK jointly prepared and submitted the applications for ethical approval and regulatory permiossions. ES also contributed to participant recruitment and data acquisition while also being responsible for the overall scientific oversight of the project. All authors participated in data interpretation, critically reviewed the manuscript, and approved the final version for publication.

## Acknowledgements

Not applicable

